# Vaccinations or Non-Pharmaceutical Interventions: Safe Reopening of Schools in England

**DOI:** 10.1101/2021.09.07.21263223

**Authors:** Carolina Cuesta-Lazaro, Arnau Quera-Bofarull, Joseph Aylett-Bullock, Bryan N. Lawrence, Kevin Fong, Miguel Icaza-Lizaola, Aidan Sedgewick, Henry Truong, Ian Vernon, Julian Williams, Christina Pagel, Frank Krauss

## Abstract

With high levels of the Delta variant of COVID-19 circulating in England during September 2021, schools are set to reopen with few school-based non-pharmaceutical interventions (NPIs). In this paper, we present simulation results obtained from the individual-based model, June, for English school opening after a prior vaccination campaign using an optimistic set of assumptions about vaccine efficacy and the likelihood of prior-reinfection. We take a scenario-based approach to modelling potential interventions to assess relative changes rather than real-world forecasts. Specifically, we assess the effects of vaccinating those aged 16-17, those aged 12-17, and not vaccinating children at all relative to only vaccinating the adult population, addressing what might have happened had the UK began teenage vaccinations earlier. Vaccinating children in the 12-15 age group would have had a significant impact on the course of the epidemic, saving thousands of lives overall in these simulations. In the absence of such a vaccination campaign our simulations show there could still be a significant positive impact on the epidemic (fewer cases, fewer deaths) by continuing NPI strategies in schools. Our analysis suggests that the best results in terms of lives saved are likely derived from a combination of the now planned vaccination campaign and NPIs in schools.

## 1 Introduction

The spread of SARS-CoV-2, and associated variants of concern (VOC), has caused significant disruption to health care systems and over 4.5 million deaths recorded worldwide as of 1st September 2021^1^, with more than 150,000 of those in England alone^2^. To combat the spread of the virus, a multitude of non-pharmaceutical mitigation strategies have been used at different times, including isolation of cases and their contacts, the closure of meeting places such as leisure venues, and the encouraging of social distancing practices. Pharmaceutical interventions, *e.g*. vaccination programmes, play a central role in negating the impact of COVID-19. They significantly reduce the likelihood of severe symptoms, hospitalisation and deaths from the original Coronavirus strain^3–5^ and they are effective against more transmissible variants, including Delta^6–8^. They are also effective in reducing the effect of infection and transmission^6,9^.

The United Kingdom (UK) began its vaccination campaign on the 8th December 2020, prioritising groups identified by the Joint Committee on Vaccination and Immunisation (JCVI), beginning with the older members of the population and those more vulnerable to severe response to the disease^10^. The protective benefit of vaccination has been manifest: relaxation of restrictions in the UK has led to a significant increase in overall case numbers, but with a much less severe increase in hospital admissions and deaths^2^. Initially, vaccines were only offered to those over the age of 18, however, from the 23rd August 2021, the offer has been extended to those over 16 years old^11^, and from the 20th September 2021, to those over 12 years old^12,13^.

While schools present a potentially important transmission route given the facilitated mixing of large numbers of households, it has been conjectured that in comparison with adults, children are both less likely to be symptomatic and less likely to be as infectious, dampening the efficacy of such transmission. Indeed, a meta-analysis^14^ finds that in studies which specifically stratify by age, children had 48% lower odds of infection compared with those over the age of 20^15^.

However, many of the clinical school studies in the UK and elsewhere were performed early in the pandemic outbreak, when testing was limited and asymptomatic cases missed, and before the advent of the Delta variant. Despite this uncertainty, one of the most commonly used non-pharmaceutical interventions (NPIs) has been the closure of schools. Indeed, by early April 2020, 188 countries, including the UK, had closed schools and more than 90% of the world’s learners were affected^14^. To mitigate the risk of school transmission while schools were open, a range of measures have been used in the UK during the pandemic thus far. In particular, following school reopening in April 2021, masks were required in secondary school classrooms, social distancing, regular testing, and a combination of bubbles, and sending home either close contacts of infected individuals or their entire bubble was used. As of September 2021, these school based NPIs have been withdrawn with only regular testing and isolation of positive cases remaining. However, unlike the adult population, most children have not yet been vaccinated.

With schools reopening in England in September 2021, assessing the potential risks of reopening is crucial. Existing UK-based studies found a wide range of impacts of school opening in comparison to community transmission, ranging from significant^16,17^ to negligible^18^, with only the latter study assessing the impact of school reopening in 2021 explicitly and none assessing their reopening in the context of the Delta variant. In this paper we focus on the relative importance of vaccination and NPIs in schools on pandemic progression in England during the latter half of 2021. We compare the influence of three different vaccination campaigns (children in the 12-15 years old cohort, the 16-17 years old cohort, and only adults) as well as the impact of continuing NPIs in schools. The influence of vaccination and the likelihood of re-infection use optimistic parameters. None of our scenarios are intended as actual forecasts, rather they are used to compare interventions.

We model these scenarios with JUNE^19^, an individual-based model which uses fine-grained geographic and demographic information and a strong focus on the details of policy interventions to describe the spread of infectious diseases. It simulates the movement of all the inhabitants of England (∼ 53 million individuals in our simulations) in a geographically resolved representation of their interactions at home, school, work, and recreation^19^. Apart from the parameters we vary to assess these scenarios, model parameters were those fitted using data from the first wave in England. This fitting allowed us to determine pre-pandemic contact intensity parameters, on top of which we can apply interventions (see original model paper^19^). We will briefly introduce reference scenarios (Section 2) and then present results in Section 3. These are contextualised and discussed in Section 4. The model and its parameters are detailed in Section 7, with details of sensitivity studies in the Supplementary Notes.

## 2 Scenarios

We use several scenarios to compare and contrast the impact of vaccination campaigns and NPIs. In these scenarios the Delta variant is assumed to be the dominant circulating virus, and some community NPIs are still in place, such as partial mask wearing and the isolation of positive cases (see Section 7). In all scenarios we assume that the vaccination campaigns were completed before the simulations began, which means none of these simulations are intended as forecasts, but rather to assess the relative effects of different interventions. Isolating vaccination in this way allows a cleaner comparison of possible interventions.

The different vaccination scenarios are:

- BASELINE: ≈ 80% of adults over 18, are fully vaccinated;
- OLDER-TEENS: Vaccinating 80% of those aged 16-17 alongside the BASELINE distribution of adult vaccinations;
- MOST-TEENS: Vaccinating 80% of those aged 12-17 alongside the BASELINE distribution of adult vaccinations.
- ALL-CHILDREN: Vaccinating 80% of all children alongside the BASELINE distribution of adult vaccinations.

Although the scenarios involving vaccinating children do not match reality in timing and practicality (e.g. vaccines have not yet been approved for their use on children younger than 12 in the UK), the exercise here is to assess the counter-factual: what would have happened had these campaigns occurred before mid-summer?

We also assess the possible impact of NPIs in the school environment under two scenarios which are variations on the BASELINE:

- CLASS-QUARANTINE: When a pupil develops symptoms, their whole classrooms isolates at home for 10 days;
- SOCIAL-SCHOOLS: Variations in the intensity of contacts between individuals in schools to mimic the effect of policies such as mask wearing, social distancing and isolation between year groups.

All simulations began from 10th July 2021 and we ran all scenarios up to 1st February 2022. The only changes we make in the simulations are the reopening of schools and universities at the beginning of September and October respectively (however, university opening has a minimal effect in our model, see Supplementary Note 5). A detailed description of the methods used with the values of key parameters is exposed in Section 7.

## 3 Results

Figure 1 shows a set of vaccination scenarios together with schools opening under the assumption of no school-specific NPIs. The left column of the upper row shows the total number of daily infections in England (for all ages, and including cases that would not be detected by testing). In the BASELINE and OLDER-TEENS scenarios we observe a significant higher increase in daily infections, with a peak in the second half of September. Here, reopening schools results in a peak approximately a factor of four (OLDER-TEENS) to five (BASELINE) times larger compared to the time just before school opening. In the scenario where most teenagers are vaccinated (MOST-TEENS), this peak in daily infections is delayed by about two weeks and less pronounced, and it is nearly entirely absent in the scenario where most children of all age groups are vaccinated (ALL-CHILDREN). The increase in infections is also reflected in the number of daily deaths in the right column, upper row, which peaks about two weeks after the spikes in the daily infections as expected, and approximately follows the same pattern of relative increases. The lower row shows the cumulative numbers of total infections and deaths during the simulations.

**Figure 1.**
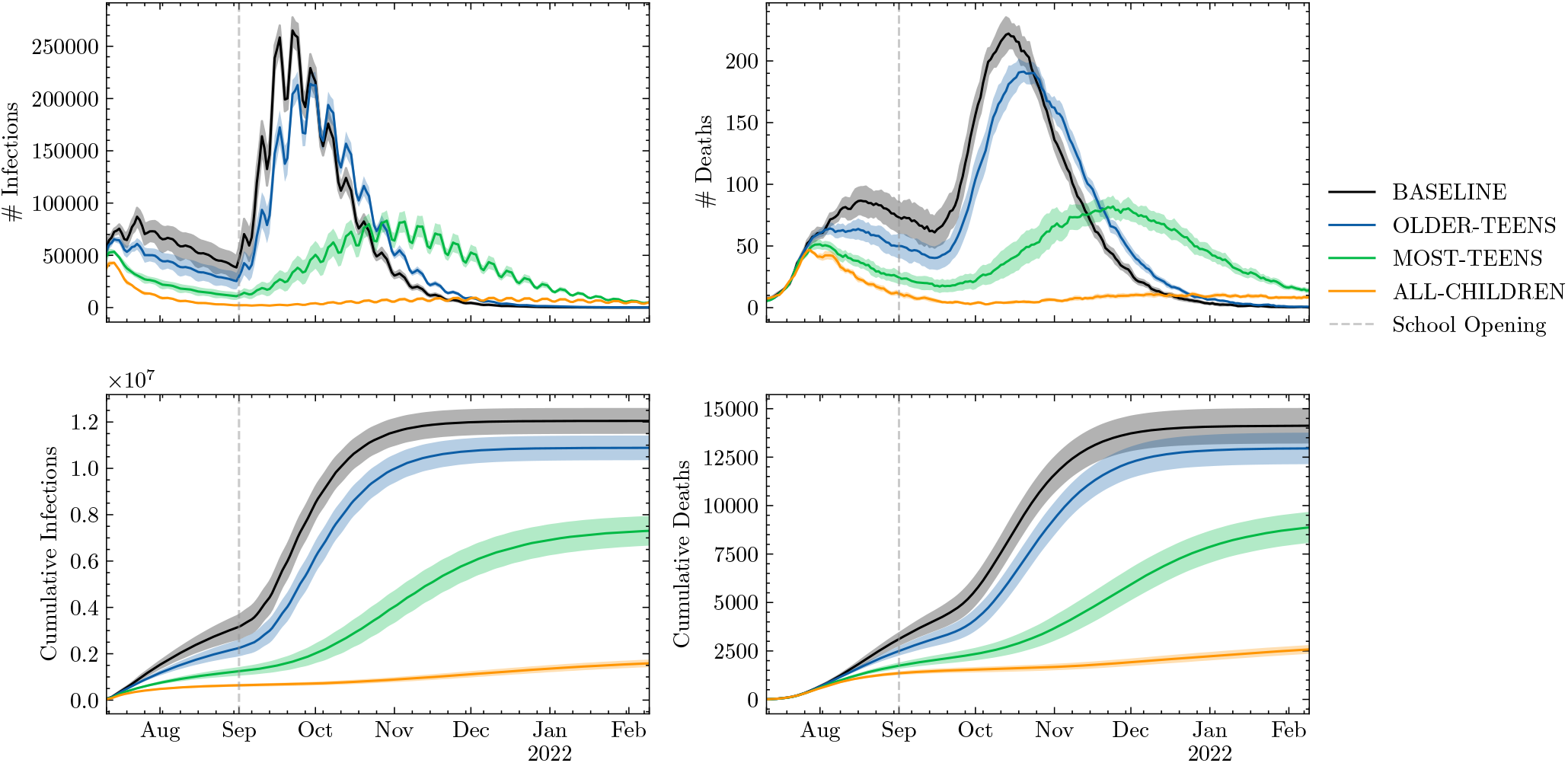
New infections (left) and deaths (right) for all ages in England per day, daily (upper) and cumulative (lower). We show four different scenarios in which ≈80% of the eligible population is vaccinated: BASELINE - only those older than 18; OLDER-TEENS - those older than 16 are vaccinated; MOST-TEENS - those older than 12 are vaccinated; and ALL CHILDREN - the entire population is eligible. Shading shows an estimate of model parametric uncertainty (discussed in Section 7).

In these simulations a successful vaccination campaign for the 16-17 year old cohort reduces the total number of infections and deaths by about 10%. This reduction is more significant when vaccinating children over the age of 12, which could have reduced the total number of infections by 40% — 7 million instead of the 12 million observed in the BASELINE scenario — and similarly drastically reduced the number of fatalities (by about 5,000 deaths). Of course, this positive outcome would have been even more pronounced in the event that vaccines for under-12s had been approved and they had been vaccinated. In the ALL-CHILDREN scenario reopening schools would have had negligible impact on infection rates and fatalities.

Figure 2 shows the impact of other interventions (NPIS) in the school environment on the daily (left column) and cumulative (right column) number of deaths. The upper row shows the impact of varying levels of contact intensity. In JUNE, this is represented as a multiplier on the parameterised intensity of social interactions in schools. Clearly, reducing interaction intensity in schools has a significant impact on the number of fatalities, and has the potential to reduce them by up to 50%. The lower row of Figure 2 shows the CLASS-QUARANTINE scenario, in which entire school classes must quarantine in the event of a symptomatic case. This policy significantly dampens the peak of the daily death curve, and the effect on the cumulative number of deaths is similar to that seen from near maximal contact intensity reduction.

**Figure 2.**
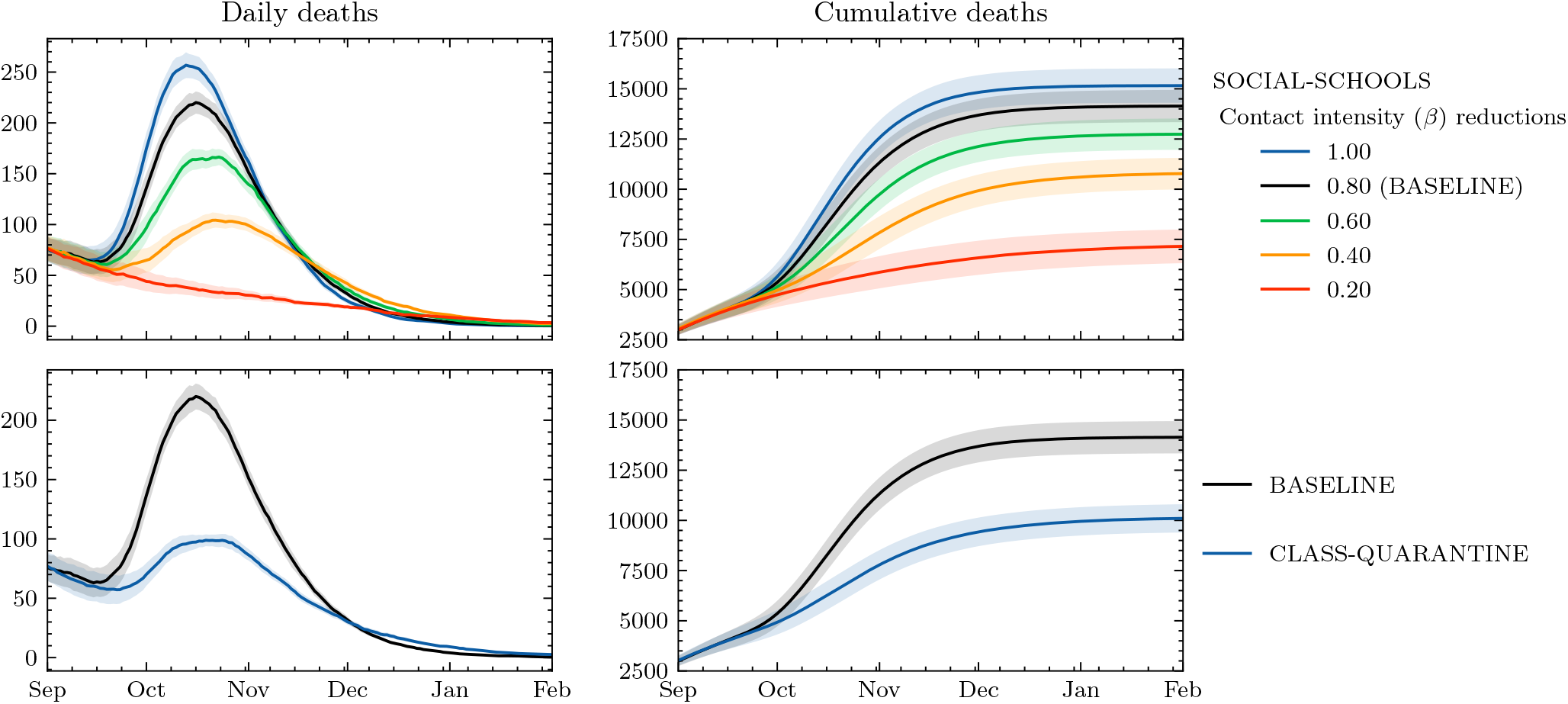
The impact of NPIs in schools on daily deaths (left panels) and cumulative deaths (right panels). Top panels: varying interaction intensities (*β* parameters) in schools (approximating social distancing, masks, and other interaction changes). Bottom panels: (re)Introducing a quarantine policy (classes sent home for 10 days if a pupil shows symptoms). Shading shows an estimate of model parametric uncertainty.

A comparison of the main vaccination and school intervention scenarios is shown in Table 1. In these scenarios the most effective interventions are a priori vaccination of MOST-TEENS, followed by the largest reductions in school contact intensity.

**Table 1.** Comparison of scenarios results respect to BASELINE. A negative value is associated with a % reduction. Ranges are the absolute values and errors from the structural uncertainty calculations.

| Scenario | $\Delta$ (infections) % | $\Delta$ (deaths) % |
| --- | --- | --- |
| BASELINE | 0 | 0 |
| OLDER-TEENS | − [10, 9] | − [9, 7] |
| MOST-TEENS | − [44, 34] | − [42, 32] |
| ALL-CHILDREN | − [98, 75] | − [93, 70] |
| CLASS-QUARANTINE | − [32, 26] | − [32, 25] |
| SOCIAL-SCHOOLS |  |  |
| β-reduction: 0.2 | − [63, 46] | − [58, 41] |
| β-reduction: 0.4 | − [31, 25] | − [27, 21] |
| β-reduction: 0.6 | − [12, 10] | − [11, 9] |
| β-reduction: 1.0 | + [8, 9] | + [6, 8] |

The age distribution of mortality in these scenarios will differ from earlier phases of the pandemic. Using JUNE results from an as yet unpublished hindcast simulation of the first phase of the epidemic, Figure 3 compares the BASELINE scenario with that first phase. Although overall significantly more deaths occurred during the first wave, asymmetries in the vaccination campaign by age group and the higher infection rates due to school reopening lead to an increased number of deaths among the younger population relative to the first wave (including ∼ 600 younger than 40, compared with ∼ 400 in the first wave). Proportionally more of these deaths occur in unvaccinated individuals (Figure 3, right), with unvaccinated people in the oldest age groups of the BASELINE scenario at almost 7 times more risk of death than a vaccinated person in their same age group (and in some cases at more than twice the risk of dying in the BASELINE simulation than in the hindcast of the first phase). It is thus likely that for the unvaccinated, or those who are immunosuppressed in whom the vaccine is less effective, the last part of 2021 will be the most risky phase of the pandemic so far.

**Figure 3.**
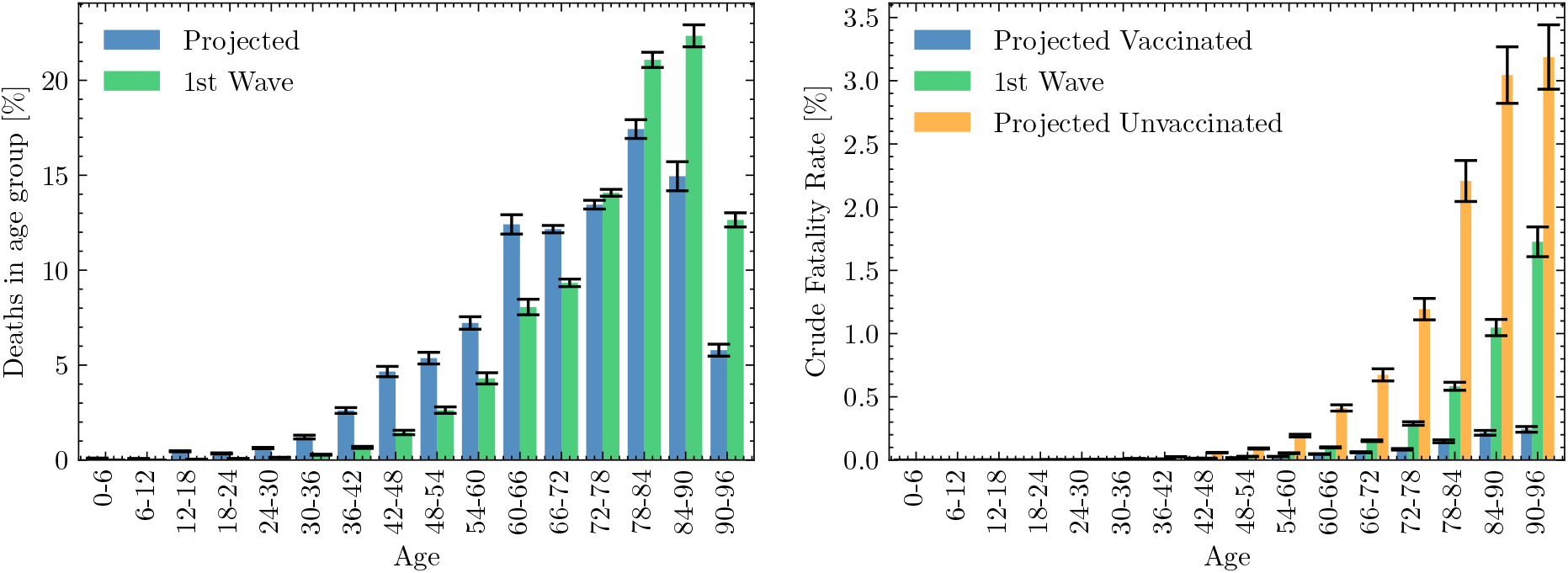
A comparison of mortality between the first-wave and that projected in the BASELINE scenario: On the left, a comparison of the distribution of all deaths in a simulation of the first wave (green) with all those seen in the BASELINE scenario (blue). On the right, the crude fatality rate seen in the BASELINE scenario for those vaccinated (orange), and unvaccinated (blue) is compared to that in a first wave simulation (green). Error bars come from comparison using the full range of beta parameter samples (see Section 7).

In all these scenarios it is clear that the impact of school re-opening is that the infections can rapidly break out of age strata (Figure 4), in line with findings from other studies^20^. It is clear that this is driven by school first infections (Figure 5), demonstrating that in this model schools present an important route for disease transmission between households. This spread of infection from schools holds true despite a large proportion of vaccinated adults, and results in significant death rates.

**Figure 4.**
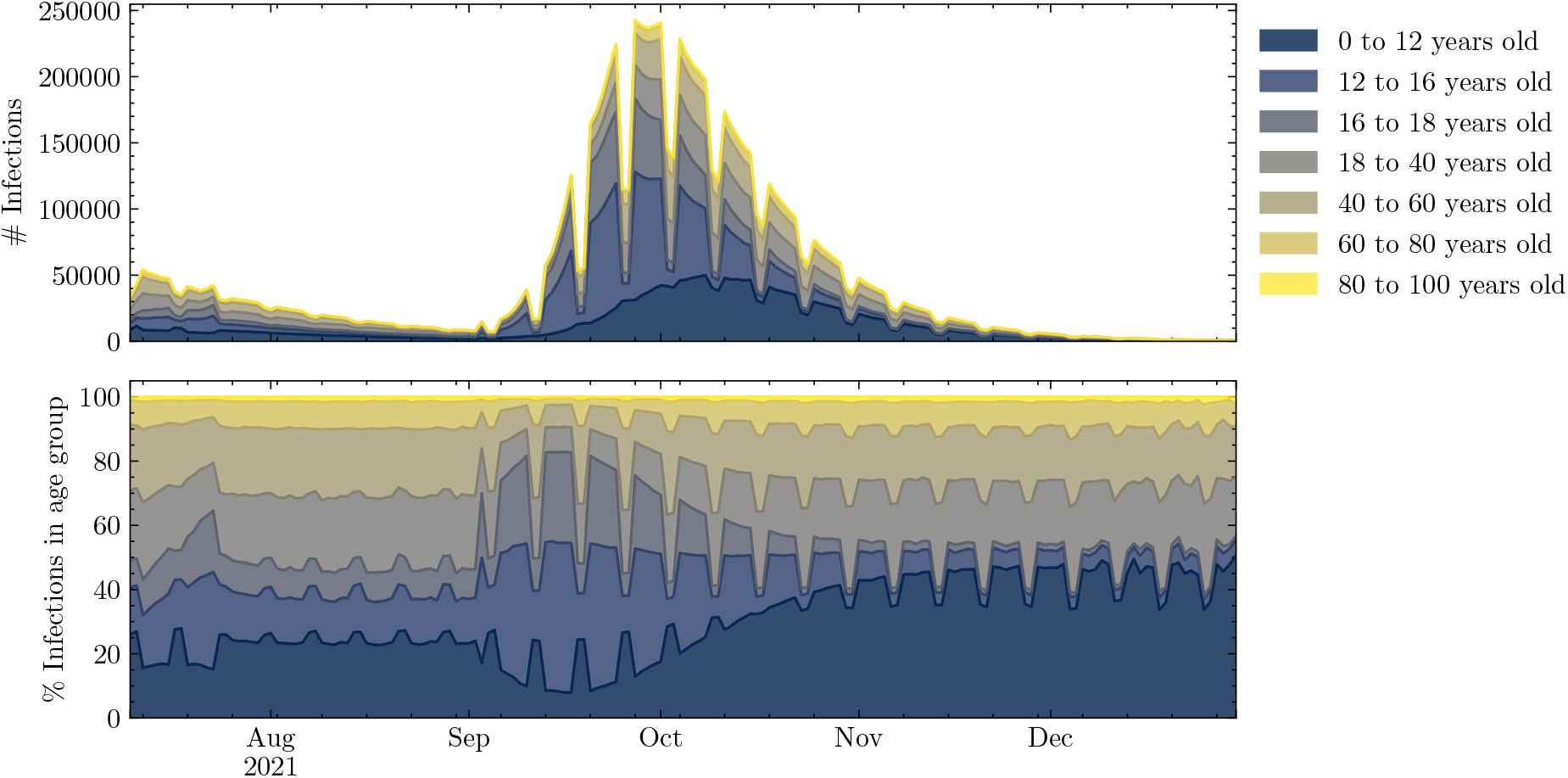
Age distribution of infections from the BASELINE scenario. Infection numbers in each age group by day (upper panel) and percentage of infections by age group (lower). Together these show increased infections among children that translate into increased infections among adults.

**Figure 5.**
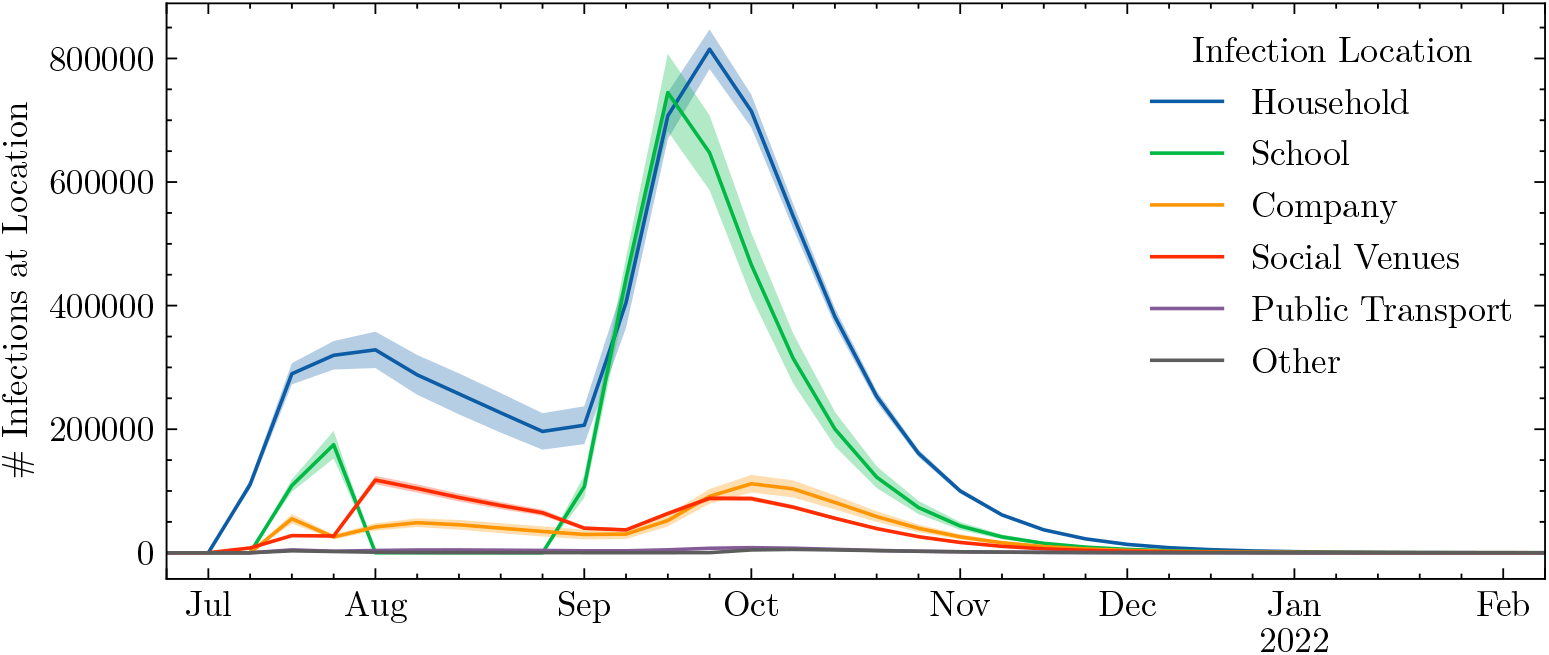
Number of infections at a given location. The peak of infections is realised first in schools, and then translated into infections in households, companies and social venues.

## 4 Discussion

We have presented results which simulate how the September-December 2021 Delta epidemic might have progressed in England under a number of scenarios.

The primary result from our comparison of vaccine scenarios is that vaccinating 80% of 12−17 year olds prior to July 2021 would have had a major effect on the epidemic progression — significantly more than just vaccinating those 16 and older or adults alone. It would have delayed the autumn peak, spread it out, and potentially resulted in thousands of lives saved.

Given that any vaccination campaign for 12-15 years old cohort must happen after schools reopen, one might ask would it then be too late? While we did not address that directly, we carried out an additional set of simulations for the OLDER-TEENS and MOST-TEEN scenarios where we initialised each with a lower percentage of a priori vaccinations (Figure 6). As well as addressing the possible impact of a lower uptake of vaccination amongst teenagers, these runs can be used to shed light on the question as to whether or not a vaccination campaign for teenagers starting after school reopening would still have a significant effect. In essence we hypothesize that a late start to a campaign will have a similar effect to an earlier start with more incomplete uptake, but even if this assumption is wrong it allows us to address the sensitivities in play.

**Figure 6.**
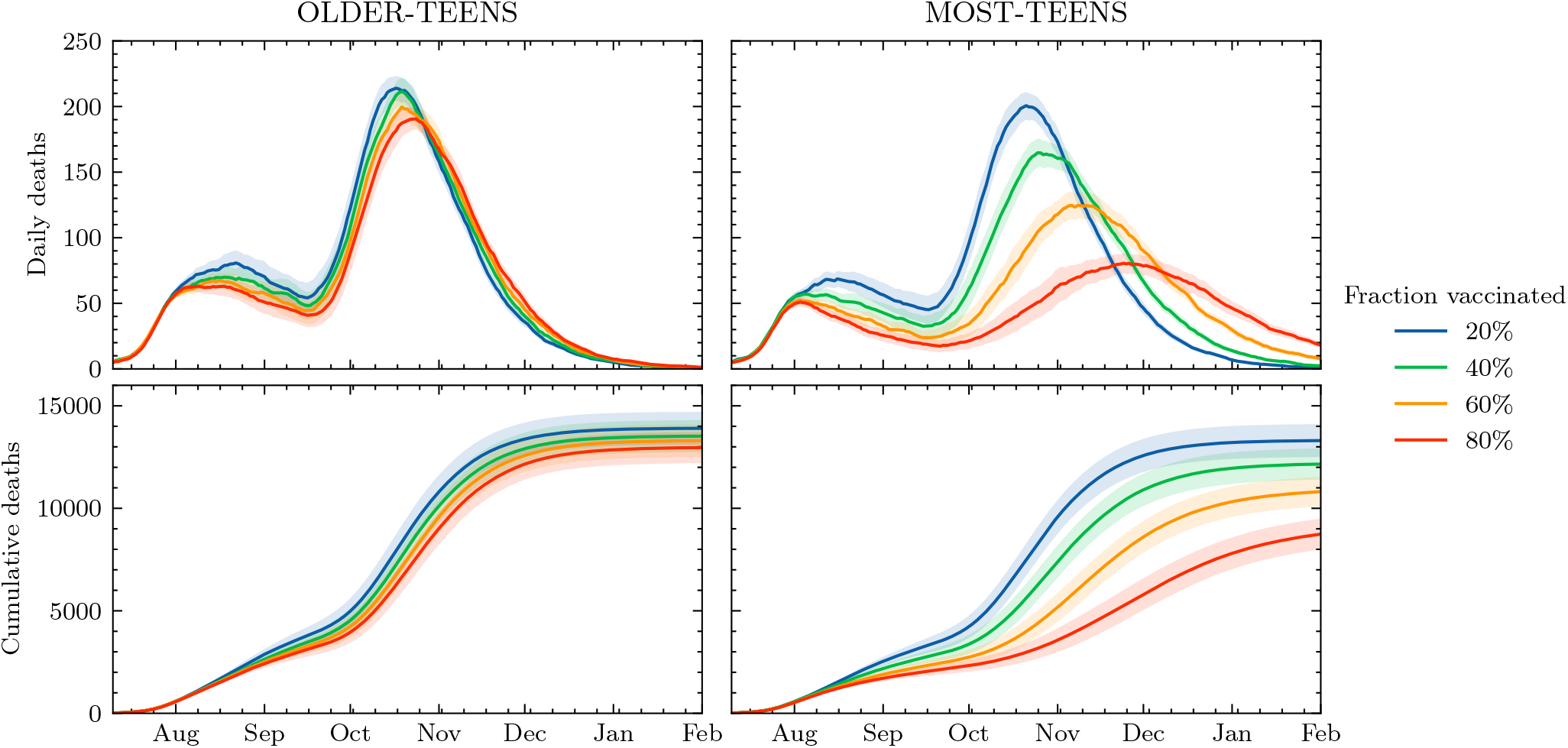
Effect on daily and cumulative deaths of changing the percentage of vaccinated children in the scenarios where we vaccinate the older teens or most teens.

Figure 6 shows that the MOST-TEENS scenario is most sensitive to a lower uptake (and hence is likely sensitive to a later start). A lower vaccination rate shifts the peak caused by school reopening to earlier dates (consistent with what one might expect from a late start) as well as increasing the peak in death rates and cumulative deaths. In the MOST-TEENS scenario doubling the vaccination rate from 40% to 80% reduces the number of deaths by 30%, but at 40% vaccination rate the death rate and death toll is not substantially reduced. While a late start could still be beneficial, in order to have a significant impact on the Delta wave, it is likely that starting a campaign to vaccinate most teenagers in September or later would need to be combined with other measures.

As an alternative or supplement to vaccination, we investigated the impact of NPIs in schools. Specifically, we have investigated two different approaches: the effect of reducing contact intensity in schools, and the impact of quarantining entire classrooms in response to symptomatic infection of one or more pupils in that class. Our results suggest the best results are achieved by reducing contact intensity in schools. Classroom quarantines were also found to be effective, comparable with the more stringent intensity reduction. In fact with enough reduction in school contact intensity, possibly also with the addition of class quarantining, the entire autumn wave is removed.

JUNE does not implement specific non-pharmaceutical interventions, it uses an “intensity multipler” over pre-pandemic contact intensity. The extent of reduction in contact intensity can be associated with a range of measures. A multiplier of 0.3-0.5 corresponds broadly to social distancing of 2 meters^21–23^, or the effect of mask wearing (depending on the type of mask worn)^21–26^. A multiplier of 0.2-0.3 has been estimated to being equivalent to a combination of mask wearing and social distancing^21–26^, with a value of 0.1-0.2 being consistent with the all these factors and increased ventilation^21–28^.

There is a legitimate concern from our results as to whether all the measures needed to reduce contact intensity to low values could or should be carried out in primary schools. It might be that the more extreme reductions in contact intensity are not achievable there, and so it may not be possible for the average contact intensity reduction to reach the values necessary to completely remove the autumn wave. We have investigated this by carrying out a set of variants of the SOCIAL-DISTANCING scenario where the contact intensity reductions were only applied in secondary schools (Figure 7). We found that a more significant effort on reducing contact intensity in secondary schools (*β* reduction factor = 0.2 in secondary schools only) would correspond roughly to a less successful overall effort (*β* reduction factor = 0.4 for all schools, Figure 1). It is also possible, given the diverse opinions in the literature, that we have over-estimated the efficacy of the achievable reductions in contact intensity. Regardless, our results suggest that an immediate imposition of at least the measures used in schools during early 2021 would be very beneficial — particularly if accompanied by stringent attention to ventilation.

**Figure 7.**
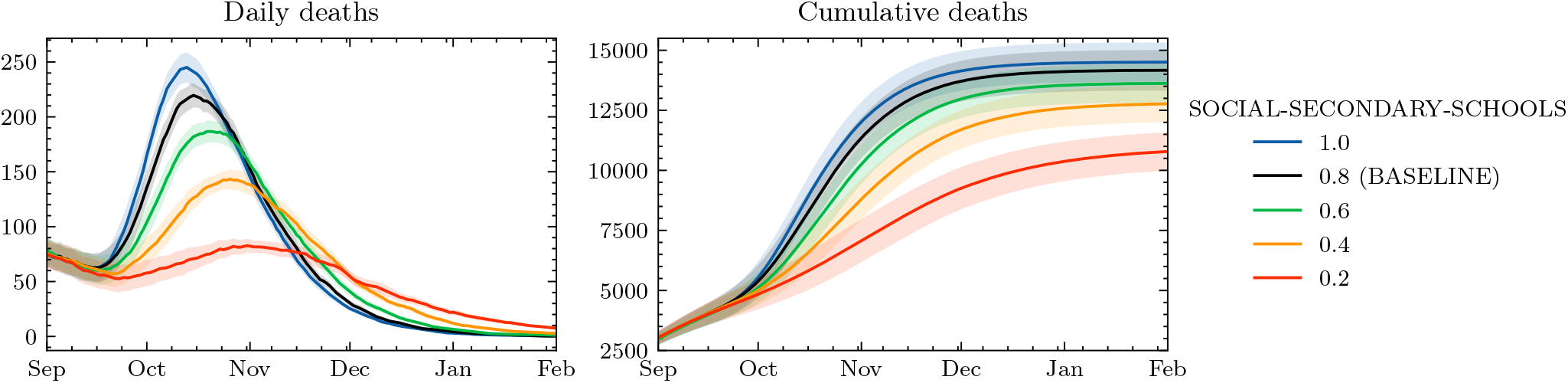
Daily deaths (left) and cumulative deaths (right) obtained while varying the contact intensity in secondary schools only, while keeping primary schools at the BASELINE value (0.8).

Our scenarios are all perturbations from the BASELINE simulation. We do not attempt to simulate a scenario with combined interventions since there is a plethora of possible combinations. However, we expect the benefit of combining vaccinations and school NPIs would be beneficial in both the simulated and real world, not least since a combined programme would both mitigate against starting the vaccination programme late and an inability to fully reduce contact in all schools, whilst at the same time it would provide a long term solution for schools to function normally.

Ours is of course not the only modelling study addressing similar issues; modelling studies focusing on the reopening of schools have been varied in their approaches and conclusions drawn. Keeling *et al*. used and age-structured compartmental model to assess the potential impact of school reopening in the UK mid-2020^16^. Their findings suggest that reopening schools without any mitigation strategies in place would likely increase the reproduction number to above one. Similar results were found by Panovska-Griffiths *et al*., who used the Covasim individual-based model^29^, and found that a second wave would likely be induced if schools were reopened in the UK alongside various other restriction relaxations^17^. The potential for negative consequences from school reopening have also been supported by similar studies conducted in other countries and settings^20,30,31^, as well as more abstract work assessing generic school settings under assumed mixing patterns^32^. Many of these works also explore possible mitigation measures such as partial school reopening, reducing classes size and increased testing, with most finding that opening earlier years is less dangerous than later years due to their reduced susceptibilities and fewer contacts.

There are studies which suggest that schools do not present such a danger. Courtemanche *et al*. found that while school closure in the United States (US) did make a positive difference to the epidemiological trajectory of the virus, their impact was much smaller than that of other measures^33^. Tatapundi *et al*. use an agent-based approach to assess the impact of partial and full school reopening in Florida, finding a less than 10% increase in cases when fully reopened in their model^34^. Minimal simulated increases in the reproduction number due to school opening have also been found in studies from Norway^35^ and Japan^36^. However, comparison of the resuse of NPIs in different countries suggest the same intervention in one country can have very different outcomes in others^37^. For the UK Sonabend *et al*.^18^ use a compartmental transmission model^38^ and include the effect of VOC and a sterilising vaccine to explore a range of scenarios for relaxing restrictions from the period June - August 2021^18^. They include school reopening in their model but find that the large scale restriction relaxations have a dominating effect on increases in the reproduction number.

Our work is most similar in methodology to that of Panovska-Griffiths et al.^17^, in assessing the most recent school reopening scenarios in the presence of various vaccination strategies. However, in comparison to theirs and other work (including Sonabend et al.^18^), our model allows more comprehensive representation of school interactions including inter- and intra-year group mixing, dividing children into classes, and greater control over possible interventions.

## 5 Limitations

Our scenarios differ from reality in several significant ways but could still be relevant to policy both now and for future epidemics. Key points of distinction include: (i) Model structural uncertainty — how well the model represents how people are distributed and interact in England; (ii) Model initialisation — how prior (non-Delta) infection and currently active (Delta) infection is distributed; and (iii) Scenario uncertainty —- How well the scenarios represent possible realities. For the purposes of this work it is the initial conditions and the representation of community vaccination and delta itself that we need to consider, other sources such as the impact of social distancing and quarantine are examined by our scenario approach, and we address some elements of model uncertainty by using an ensemble of simulations (see Methods, Section 7).

For vaccination, the most important deviation from reality is that in all our scenarios all vaccinations were completed prior to the simulation starting 10th July. (While the JUNE framework does offer a time-dependent implementation of vaccination distribution, it was not utilised in this study.) In reality at this date only 64% of the adult population was fully vaccinated, but by the end of August was nearer 80%^39^. If our initial conditions were fully faithful to reality this difference would likely to lead to an underestimate in cases and deaths in comparison to data. We do see that in the first few months of our simulations, but there are also a variety of other reasons why this might be (see Supplementary Note 4).

Conversely, if we have under-estimated the prior prevalence of the virus then this would result in fewer deaths. We tested the sensitivity of our model to prior infections and found that, while that does indeed lower deaths in our simulation, the qualitative shape of the epidemic progression remains similar and we might expect our comparative results to remain similar. We do not attempt to correct the BASELINE scenario since fixing this discrepancy might simply compensate for other issues with our initialisation and we do not claim that BASELINE is a forecast, simply that it represents a plausible reality.

An additional source of uncertainty around our representation of Delta is how much more infective it is than wild-type Covid-19 (and the Alpha variant which infected many in the English wave-2)? We investigate our sensitivity to this parameter (Figure S1) and find that by the beginning of September most values of infectivity give similar qualitative and quantitative results. Importantly, all simulations show an increase in both infection and deaths due to schools re-opening.

Together our set of assumptions around infectivity, low susceptibility following prior infection with other variants, no re-infection following infection by Delta, and a time-dependent vaccine efficiency (no waning immunity) can be seen as conservative. Our sensitivity analysis suggests that these could also be thought of as optimistic, it is possible that these artificial scenarios could underestimate the progression of the real epidemic. In particular, should our assumptions around waning immunity and lack of re-infection by Delta be incorrect, the epidemic could not only be larger than simulated, it could last longer. However, even if so, we believe our conclusions around the relative impact of interventions would be robust.

## 6 Conclusion

In this paper, we have used the individual-based model, JUNE, to simulate the spread of COVID-19 infections after reopening of schools in England from September 2021. Taking a scenario-based approach, we have highlighted the strong possibility that this induces a significant peak in the number of daily infections, corresponding to a greater number of deaths. Through the use of vaccination programmes targeting young people, these negative epidemiological consequences may have been mitigated. Had the UK vaccinated most children over the age of 12, our findings suggest that the impact of reopening schools could have been reduced by a factor of 2, with vaccinating all children removing the risk of an autumn wave entirely. However, we also find that vaccinating only those children aged 16-17 would have a relatively small effect.

In addition, we explored a range of NPIs to help mitigate viral spread. Specifically we find reducing contact intensity as much as possible (such as combining mask wearing, social distancing, and increased ventilation in classrooms) could limit the spread of the virus to the point where it would be indistinguishable from the case of vaccinating a large fraction of children. Correspondingly, less stringent measures, such as mask wearing or social distancing alone, will not be a efficient in reducing the cumulative number of deaths.

A careful analysis of model uncertainties was carried out, along with a sensitivity analysis, to test the robustness of our overall results; this analysis does not preclude the possibility that we could be under-estimating the progression of a real epidemic. While our simulations are not intended as forecasts, they suggest that the best outcome for England, in terms of cumulative deaths due to Delta during autumn 2021, would occur if a vaccination programme was begun as soon as possible for children over the age of 12 and that a mixture of NPIs should be put in place in schools while that programme is rolled out.

## 7 Methods

We use JUNE^19^, an agent-based modelling framework developed to simulate the spread of infectious diseases with a fine-grained geographic and demographic resolution. In this study, we follow the setup presented in previous work^19^, although the framework is designed to be adaptable to other settings as well^20^. We simulate the movement of the 53 million inhabitants of England, and the spread of the disease in a geographically resolved representation of their interactions at home, school, work,and recreation.

The ‘world’ in which the agents move (where they work and live etc.) uses census data from 2011, and statistical representations of their distribution into geographically dispersed households, workplaces, schools etc. (including, at appropriate times, transport units to explicitly model both local and long-distance commuting). The frequency and intensity of interactions is controlled by prescribed contact matrices and intensity parameters (from now on βs), and infections are transmitted between individuals in contact using a representation of the infection agent (in this case Delta) and the susceptibility of those in contact with an infection person. Susceptibility is a function of age, vaccination status, and prior infection.

In the remainder of this section we discuss how we represent the Delta variant, how the model was initialised, how we characterise model uncertainty, and how we implement the scenarios in this model (and in particular how we represent policies and social distancing in schools). Together these each represent a key source of uncertainty in interpreting our results (uncertainty is further discussed in Supplementary Note 4).

### 7.1 Representing Delta

The COVID-19 variant B.1.617.2 (Delta), which was first detected in India, has overtaken the B.1.1.7 (Alpha) variant which previously dominated cases in the UK. There is now strong evidence suggesting higher transmissibility compared to that of the first wave virus (wild-type)^40^. We represent variants in JUNE by introducing multiple different types of infection into the model and using a multiplicative factor for each type of transmission over wild-type. For these simulations we only use one infectious variant in the model, and make it twice as infective as wild-type. We have investigated other possible values of this multiplicative factor (Supplementary Note 2.1) and conclude that this factor of two generates plausible levels of deaths and infection.

Given that the probability for re-infection with a new variant is estimated to be low^41^, we assume that individuals previously infected by any COVID-19 variant are only 2% susceptible to any variant of the virus, compared to those that have not experienced a previous infection. We have investigated our sensitivity to this factor — not surprisingly increasing susceptibility has the effect of increasing the scale of the epidemic but most of the effect is before school re-opening (Figure S2).

### 7.2 Initial Conditions

The primary concern in initialising these runs is to establish an appropriate distribution of prior infections, currently active infections, and prior vaccination (according to the BASELINE scenario).

By the beginning of the simulation we assume that 25% of the population already had some variant of the virus^42,43^. We distribute these previous infections weighted by the number of positive cases observed by region and age (Figure S4).

For current infection we randomly distribute 577,000 infections across the full population of England using estimates from the 10th July 2021 from the ONS Infection Survey^44^. These are also weighted by the distribution of cases by age and region during that week. Although our initialisation weights cases by age and region, it does not take into account the fact that infections grow in local clusters. However, within a few weeks, when millions are infected, the original distribution is likely not relevant to the results we present.

All vaccinated individuals are assumed fully vaccinated (i.e. we do not distinguish between single and double dosed) before the simulations start. Vaccines are distributed according to age and location using ONS data^45^ for the 10th of August, resulting in ≈ 80% of the simulated adult population being vaccinated. Although three vaccines are being distributed in the UK, we only include vaccinations with AstraZeneca and Pfizer vaccines, since these dominate usage and are the main subjects of most prior studies^43^. We distribute vaccine types among the population according to their age using estimates from Scottish data^46^. The resulting distribution of total and AZ vaccinations by age are shown in Figure S5.

Throughout all simulations we assume that there is no waning immunity (neither from vaccine or previous infection) and that children aged between 0 and 12 are half as susceptible as adults of being infected^15^. Since these assumptions are shared among different scenarios, we still capture variations due to different group of the population not being vaccinated.

### 7.3 Vaccines

We assume that vaccines can protect individuals through two independent mechanisms: (i) by reducing their susceptibility of being infected; and (ii) by reducing their probability of developing a severe condition after infection.

Implementing (i) is not straightforward, since the measured vaccine efficacy against infection, *VE*_infection_, is dependent on the population dynamics. A naive assumption would be to set the susceptibility of all vaccinated individuals to a value of (1−*VE*_infection_), but this does not guarantee recovering the measured efficacy when performing a population survey, which is calculated as

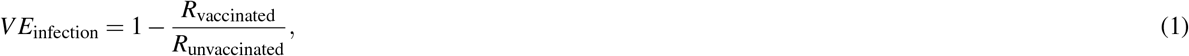

where *R* represents the risk of infection for either the vaccinated or the unvaccinated adult population, i.e., the number of infections divided by the population size. To circumvent this, given a value of *VE*_infection_ measured in a population survey, we set the susceptibility of vaccinated individuals to the value that reproduces the measured *VE*_infection_ when performing a mock survey with the same characteristics on the JUNE population. We refer to Supplementary Note 3 for a detailed explanation of the process. The resulting *VE*_infection_ values for the AstraZeneca (AZ) and Pfizer vaccines are show in Table 2.

**Table 2.**
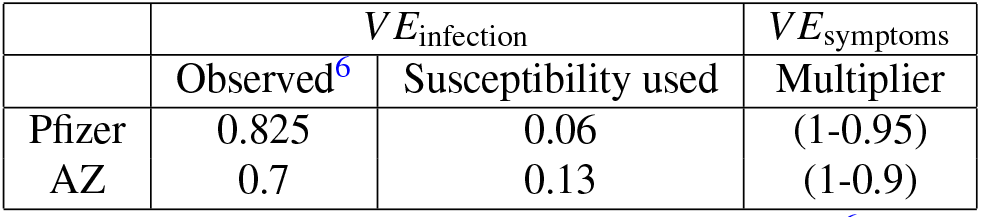
Vaccine efficacy values: Observed values from Pouwels *et al*.^6^ are implemented by setting infection susceptibility of vaccinated individuals for different vaccines and a multiplicative factor is used to achieve symptomatic efficacy.

| | $VE_{\text{infection}}$ | | $VE_{\text{symptoms}}$ |
| --- | --- | --- | --- |
|  | Observed<sup>6</sup> | Susceptibility used | Multiplier |
| Pfizer | 0.825 | 0.06 | (1-0.95) |
| AZ | 0.7 | 0.13 | (1-0.9) |

We implement (ii) for each infected individual by multiplying the probability of that they develop severe symptoms by (1 −*VE*_symptoms_), where *VE*_symptoms_ is the vaccine efficacy against symptoms, therefore reducing the probability of death and hospitalisation by the same number (Table 2).

Although there is also evidence that vaccines lose efficacy against the Delta variant^6–8^, this seems mainly to arise from a fall in the sterilising efficacy (*VE*_infection_). We investigate the impact of changing this value in Supplementary Note 2 but find the value we use provides the best agreement with reality.

### 7.4 Model Uncertainty

The model is defined by 15 free parameters: 14 *β* parameters corresponding to the contact intensities in each location, and 1 parameter (*α*_p_) that models the relative intensity of physical contacts to non-physical contacts (the original JUNE paper^19^ provides a detailed explanation on how contacts in locations are parameterised). These parameters need to be fitted against data on previous deaths and hospitalisations.

We began by setting model parameters selected from an extensive exploration of the model’s 15 dimensional parameter space using latin hypercubes which were then refined using Bayes linear emulation and history matching methodologies^47–49^ following the original JUNE methodology^19^.

The resulting sets of *β* parameters included many with plausible results, and so we sampled 8 well spaced parameter combinations from this plausible parameter region exploring the extremities of the region. Figure 8 shows a comparison of those 8 JUNE simulations to the observed daily death data.

**Figure 8.**
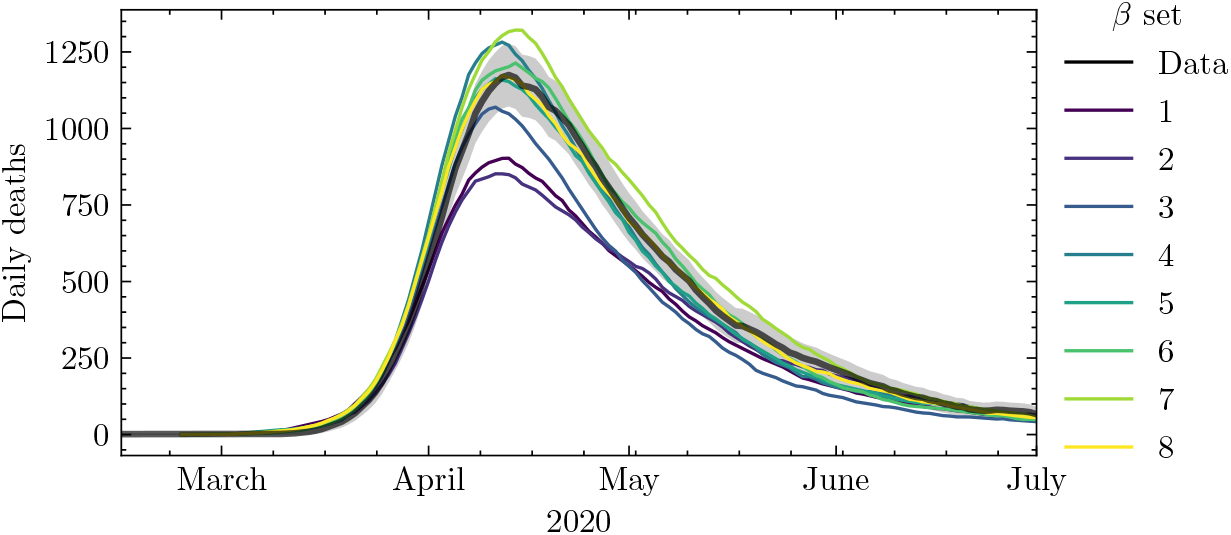
Daily deaths produced by the selected 8 JUNE *β* sets compared to data. The dark shading region corresponds to a 3-sigma error in the data. Both the data and the JUNE curves have been smoothed by averaging over a time period of 7 days.

We treat the 8 sets of *β* parameters as estimates of the model parametric uncertainty and apply them in to the BASELINE, vaccination and NPI scenarios. With sensitivity experiments (Supplementary Note 2), we run in excess of 150 simulations (Table S1).

Despite all the simulations, there still key sources of structural uncertainty: for example, we do not have representation of “holidays” and the associated mixing following domestic travel (let alone international travel). We do not model nosocomial infection, despite that being a significant source of infection early on in the pandemic^50^. Undoubtedly these sort of missing interactions will effect direct comparisons between JUNE simulations and the real world, but they are likely to be less important for comparing intervention scenarios such as ours.

### 7.5 Policies

Policies are used to control interventions in JUNE and can parameterise changes in social behaviour, as well as the total or partial closure of certain venues (such as schools over the holiday period, or leisure venues to mirror government policy). More details on the use and implementation of policies can be found in the original model paper^19^. Here we briefly discuss the policies used across all scenarios.

At the time of writing, quarantining/isolation policies are still in place in the UK and we assume they are for the duration of our scenarios. We assume that symptomatic individuals, and members of their household, quarantine in their homes with a compliance rate of 79% based on ONS studies^51^. We only model COVID-19 circulation and so all symptomatic individuals are positive cases by definition, and we do not currently account for the testing of asymptomatic positive cases. As of the 16th August, the government announced that fully vaccinated individuals do not have to quarantine if they live with someone who has tested positive but do not present symptoms^52^. This has also been implemented into the model such that this policy activates on this date. Severely symptomatic individuals are assumed to be too unwell to leave their homes and effectively quarantine in the model.

There are still many individuals and organisations exercising caution beyond the required government regulations. Recent surveys suggest that 12% of people are still avoiding going to work in person^53^, with 5% on furlough^54^, and key workers still attending work as normal (which make up 19% of the working population^55^). Masks are still worn by many individuals and we assume that this is done in during commuting and going to grocery stores with a compliance level of 69% derived from YouGov surveys^56^. These factors are all represented in the model and assumed to stay constant throughout the simulations. However we do vary one other parameter during the simulation: As OpenTable data^57^ shows that more people are going to restaurants relative to 2019 data (at the level of 20-30%, we assume this to be the case up until the reopening of schools, after which these values are restored to pre-pandemic estimates.

The *β* multipliers discussed in Section 3 can be used to implement NPIs such as social distancing and other contact intensity changes. We assume some residual degree of caution in the population in certain venues ranging from minimal distancing (*β* multiplier of 0.9) to more regulated 1m+ distancing (*β* multiplier of 0.7^21^). These multipliers are implemented separately to those of the mask wearing policies. We assume minimal distancing in pubs, commuting units (carriages), gyms and visiting other households. Some distancing is assumed in grocery stores, care homes, companies, universities and schools, with more enforced distancing in cinemas. A complete set of parameters is included in the code release.

All venues are considered as open throughout the simulations, with the exception of schools, which open on the 2nd September, and universities, which open on the 28th September.

## Data Availability

Data will be made availble on Zenodo or a similar repository before publication.

## Data Availability

The bulk of the simulation results used for this study are available on Zenodo – https://zenodo.org/record/5520923 (some higher volume data is not included, but available from the authors). We publish in https://github.com/IDAS-Durham/june_vaccines the code to produce the figures appearing in this paper from that data.

## Code Availability

This work used the version 1.1.1 of JUNE (https://github.com/IDAS-Durham/JUNE)^58^.

## Ethics

Ethics approval was not required for this study as the data we use is open. We use the Health Research Authority (HRA) decision tool and consulted HRA formally who provided formal acknowledgment of our exemption.

## Acknowledgements

J.A.B., C.C.L., A.Q.B. thank the STFC-CDT DDIS for their support through grant ST/P006744/1. F.K. gratefully acknowledges funding as a Royal Society Wolfson Research Fellow. We gratefully acknowledge the generous provision of computing time on the Hartree and JASMIN facilities. The bulk of this work used the DiRAC@Durham facility managed by the Institute for Computational Cosmology on behalf of the STFC DiRAC HPC Facility (www.dirac.ac.uk). The equipment was funded by BEIS capital funding via STFC capital grants ST/K00042X/1, ST/P002293/1, ST/R002371/1 and ST/S002502/1, Durham University and STFC operations grant ST/R000832/1. DiRAC, Hartree and JASMIN are part of the National e-Infrastructure.

## Author contributions statement

C.C.L., A.Q.B., J.A.B., B.N.L., K.F. conceived the experiment(s); C.C.L., A.Q.B. ran the experiment(s); C.C.L., A.Q.B., J.A.B., B.N.L., M.I.L, A.S., H.T., I.V., J.W., F.K. developed the original model; C.C.L., A.Q.B., J.A.B. adjusted the model for this work; C.C.L., A.Q.B., J.A.B., B.N.L. analysed the results; C.C.L., A.Q.B., J.A.B., B.N.L., C.P., F.K. wrote the manuscript. All authors reviewed the manuscript.

## Supplementary Note 1: Simulations and Parameters

The complete list of simulations and parameters used for this study are shown in Table S1. We estimate that these simulations used 400,000 HPC core-hours and would have been responsible for about 4, 000 kg of CO_2_ emissions - equivalent to about 40, 000km of flying for one passenger^59^.

**Table S1.** The complete set of additional simulations which we compare to the BASELINE scenario (for which there were also 8 simulation spanning the possible *β* values). The upper set are discussed in the main body of the paper, the others are discussed in these supplementary notes.

| Parameter | Values varied | $\beta$ set | #runs | Figure |
| --- | --- | --- | --- | --- |
| Age threshold of vaccinated children | 0, 12, 16 | 1-8 | 24 | Figure 1 |
| All school's contact intensity reduction | 0.2, 0.4, 0.6, 1.0 | 1-8 | 32 | Figure 2 |
| School's quarantine | True | 1-8 | 8 | Figure 2 |
| Children vaccination rates | 20%, 40%, 60% | 1-8 | 48 | Figure 6 |
| Secondary school's contact intensity reduction | 0.2, 0.4, 0.6, 1.0 | 1-8 | 32 | Figure 7 |
| Delta infectivity | (1.5 – 2.8) | 8 | 14 | Figure S1 |
| Previously infected susceptibility | (0.05 – 0.5) | 8 | 6 | Figure S2 |
| % of previously infected | All (-5, +5, +10)%<br>Children (+10,+20,+30,+40)% | 8 | 7 | Figure S2 |
| Vaccination sterilisation efficacy | (0.8 – 1.0) | 8 | 40 | Figure S3 |

## Supplementary Note 2: Sensitivity Analysis

The core scenarios are underpinned by several sensitivity analyses. To mitigate computational cost, the sensitivity analysis is performed with a single set of *β* values, corresponding to run number 8 in Figure 3. We refer to the vaccination scenarios that result from this set of betas and the parameters assumed in the main text as the” representative” (REPR) scenario, since it produces the closest results to the mean of all 8 simulations in all scenarios.

### 2.1: Delta infectivity

**Figure S1.**
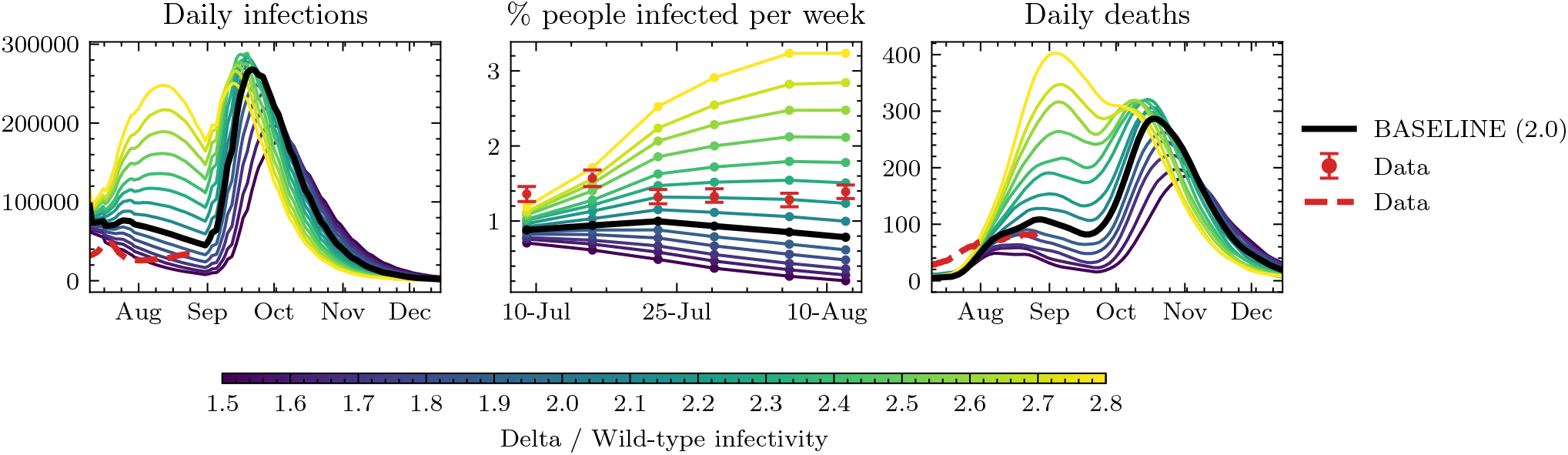
Comparison of different runs changing the infectivity of the Delta variant respect to the wild type. The infectivity is colour coded from light blue (1.5) to purple (2.8), with our baseline seating at 2.0. Left panel: Daily infections compared to cases reported^44^. Middle panel: Percentage of people respect to the total population infected each week during the months of July and August. The data corresponds to an ONS estimate^44^. Right panel: Daily deaths compared to the ones reported by PHE^60^. All daily data has been smoothed by averaging over a period of 7 days.

The Delta variant is estimated to be (40− 60)% more infectious than the Alpha variant^61^, which in turn is also estimated to be (40−70)% more infectious than the Wild-type SARS-CoV-2 that circulated in England during the first wave^62^. Consequently, we explore the sensitivity of our baseline scenario on the infectivity of the Delta variant by varying its value in the range 1.5 − 2.8. In Figure S1, we show 3 comparisons to 3 different data sources: (i) our simulated daily infections to reported ONS cases^44^, (ii) the % of people infected per week to the ONS modelled estimate^44^, and (iii) our resulting daily deaths to PHE data^60^. As we can see, no single value of the Delta infectivity is a good match to the three data sources. Apart from model uncertainty, these deviations are expected for two reasons: reported cases are a lower bound to the number of total infections, since not all of them are detected (especially asymptomatic cases), and our model initialisation routine may not be representative of the amount of active infections at the beginning of the simulation so we expect small deviations especially in the deaths data where timings are more sensitive. Overall, the position of the peak after schools reopen is not very sensitive to the Delta infectivity, and we choose an infectivity value of 2.0 since it is a good compromise in the comparison to data.

### 2.2: Previous infection

For the main text simulations we assumed that previously infected individuals have a susceptibility of 0.02 to be reinfected with any variant. Even though this assumption is likely reasonable^41^, we do not model immunity waning and so we also explore the sensitivity of the results in the amount of people with immunity caused by previous infection, and its corresponding susceptibility to reinfection. The results are show in Figure S2, where we observe that despite the fact that the overall number of infections is sensitive to varying these parameters, the position of the daily deaths peak is not, so the time of highest pressure in medical facilities is the same in all scenarios.

We have not yet explored the impact of re-infection by Delta, or of waning immunity from vaccination. We might expect either of those to prolong the epidemic through winter.

Finally, we assess the sensitivity of our results to the percent of previously infected individuals. We vary the percentage of previously infected by age, shown in Figure S4 by −5%, +5%, +10%, in each age group, and by +10%, +20%, +30%, +40% for children only. Although the overall number of deaths varies quantitatively, reopening schools effectively causes an infection peak across all variations, albeit a very damped one for the extreme case where we have lots of previous infection among children.

**Figure S2.**
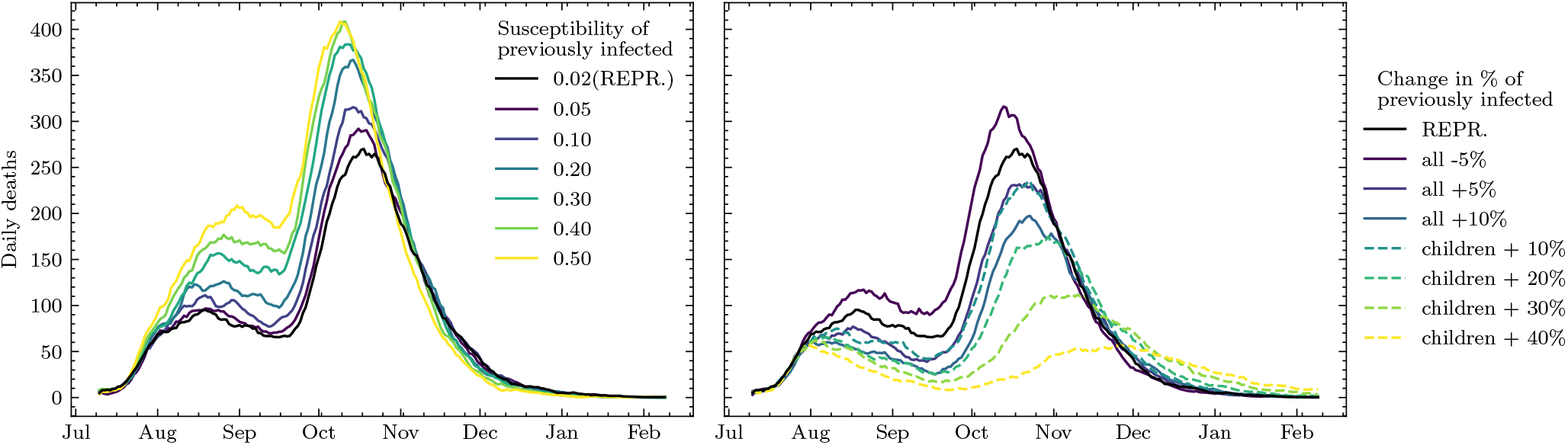
Results of simulations exploring the dependence of daily deaths on the susceptibility to reinfection of individuals infected before our simulations start (top) and the percentage of people who had been infected before our simulations start (bottom). We vary the % of the previously infected individuals as a flat change to each age group (refered as “all”), and a flat change over children only.

## Supplementary Note 3: Vaccine efficacy against infection

As explained in Section 7.3, we adjust the vaccinated agent’s susceptibility of being infected so that we obtain the right vaccine efficacy against infection. To assess vaccine efficacy against infection in JUNE we compute

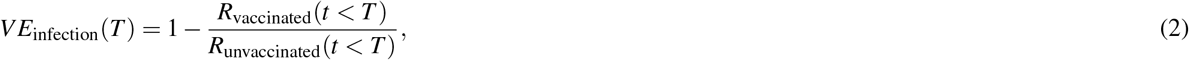

at a given time *T*, by computing the percent of infected in the vaccinated and the unvaccinated population up to that time. The vaccine efficacy is therefore a function of time, although we find that it converges to a constant value. Note that we include only adults in the assessment, since unvaccinated children could bias our measurement of vaccine efficacy. In Figure S3, we show the vaccine efficacy for both AstraZeneca and Pfizer found by varying the susceptibility set for a vaccinated individual. The susceptibilities used for all scenarios which result from this procedure are shown in Table 2.

**Figure S3.**
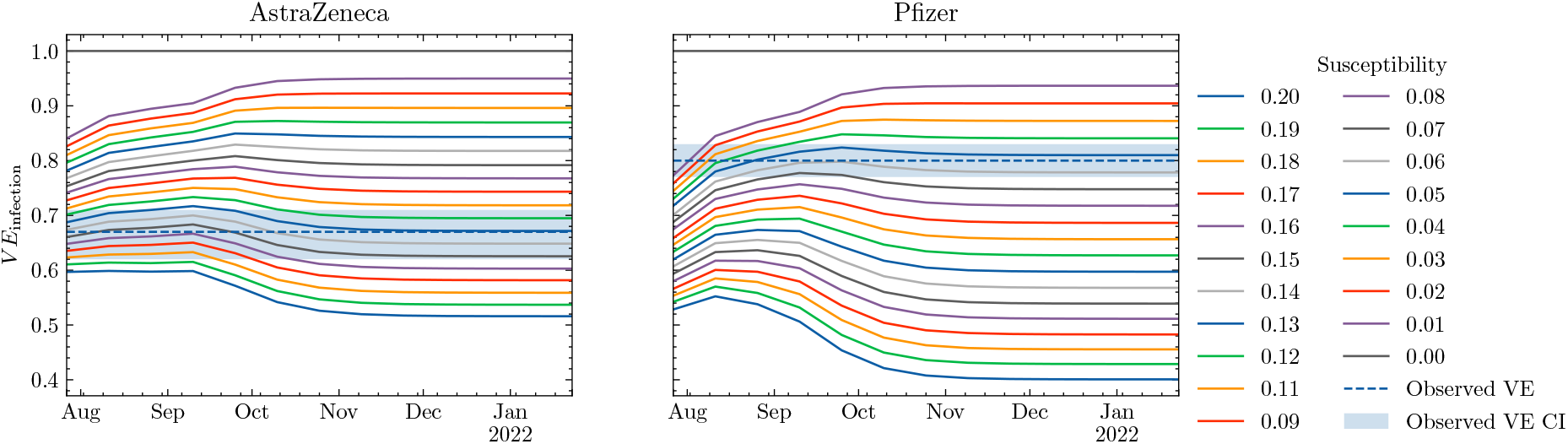
Assessed vaccine efficacy against infection in JUNE (coloured lines) as a function of the susceptibility of the vaccinated individuals. Solid bands show observed vaccine efficacy values inside the confidence interval estimated from data by Pouwels *et al*.^6^

## Supplementary Note 4: Sources of Uncertainty

Our scenarios differ from reality in several significant ways as summarised in Section 5. Here we provide some additional detail to that discussion.

For vaccination, the most important deviation from reality is that in all our scenarios all vaccinations were completed prior to the simulation starting 10th July. In reality at this date only 64% of the adult population was fully vaccinated, but by the end of August was nearer 80%^39^. If our initial conditions were fully faithful to reality this difference would likely to lead to an underestimate in cases and deaths in comparison to data. Figure S1 shows that this the case for the BASELINE scenario (at least for the ONS data and deaths, if not daily case data) regardless of how much more infective we make the Delta variant than wild-type. However the same figure shows that by late August (the last available data) while we are still underestimating cases, we are overestimating deaths. The most likely reason for this is that we have under-estimated the prevalence of prior infection and/or our statistical allocation of prior infection has not infected the right people. However there are many other factors that could play a role here: including the health index might be different to that of the first wave (e.g. due to delta variant itself and/or hospitals getting better waning immunity; people’s behaviour could be changing in ways we have not represented etc.

The hypothesis that we have under-estimate the prior prevalence can be investigated. Figure S2 shows the effect of increasing the percentage previously infected would indeed drop the numbers dying into the observed range, but it also shows that the qualitative shape of the epidemic is unchanged. We do not attempt to correct the BASELINE scenario since fixing this discrepancy might simply compensate for other issues with our initialisation and we do not claim that BASELINE is a forecast, simply that it represents a plausible reality. The important message is that the simulations are qualitatively doing the right thing.

A key source of uncertainty around our representation of Delta is how much more infective it is than wild-type Covid-19 (and the Alpha variant which infected many in the English wave-2)? We investigate our sensitivity to this parameter in Figure S1. Our main conclusion is that the impact of uncertainty in this factor primarily manifests itself during the early part of the simulation following initialisation - primarily when we think we could have too many susceptible individuals (too little prior infection). By the beginning of September most values of infectivity give similar qualitative and quantitative results. We have used a value of 2.0 (Delta twice as infective as wild-type Covid-19) as a good compromise to fitting the available data.

## Supplementary Note 5: Universities

Universities are opened on the 28th of September in all scenarios, however this has minimal effect in our simulations as most of the student population are either fully vaccinated and/or have been previously infected. Following our previous work^19^, students are assumed to be living in the same households (often communal) for the whole simulation period (since there is no available data on student in-term and out-of-term addresses).

## Supplementary Note 6: Initial Conditions

This note provides additional information about the distribution of prior infections and vaccinations used for the initial conditions of all scenarios:

- Prior infection is distributed by age and region as shown in Figure S4.
- The distribution of vaccination in the BASELINE scenario (those over 18 only) is shown in figure S5.

**Figure S4.**
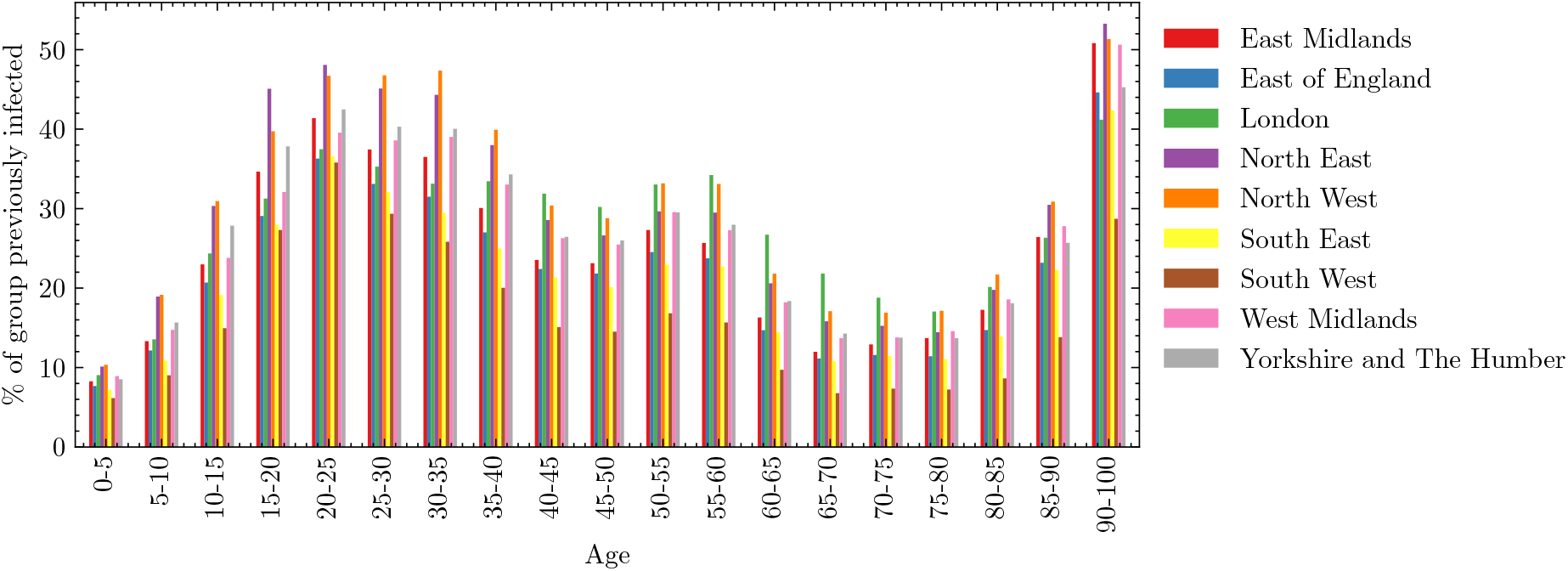
Distribution of previous infections by region and age, estimated from total cases detected in England from the begining of the pandemic.

**Figure S5.**
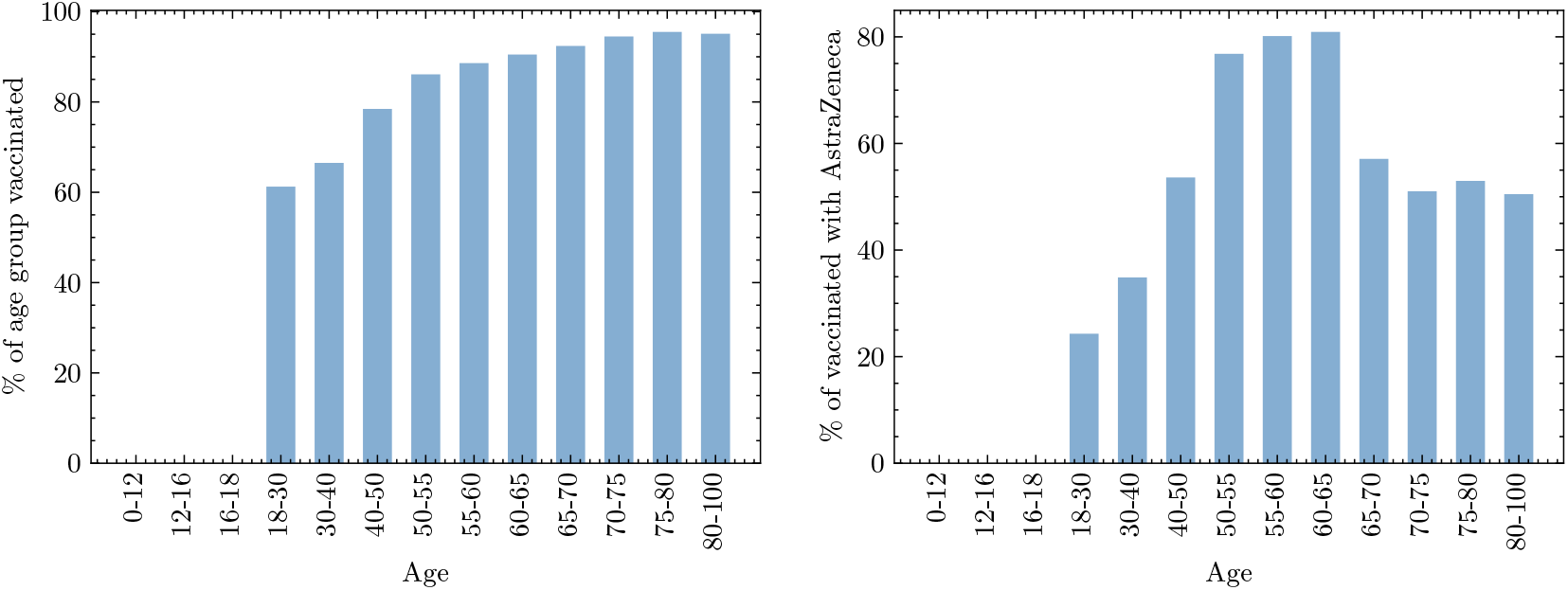
Left, percent of the population vaccinated as a function of age in the BASELINE simulation. Right, percent of those vaccinated that received AstraZeneca.

## References

1. WHO Coronavirus (COVID-19) Dashboard. https://covid19.who.int. Accessed: 2021-09-02.

2. Coronavirus (COVID-19) latest insights. https://www.ons.gov.uk/peoplepopulationandcommunity/healthandsocialcare/conditionsanddiseases/articles/coronaviruscovid19/latestinsights. Accessed: 2021-09-04.

3. Polack, F. P. et al. Safety and efficacy of the bnt162b2 mrna covid-19 vaccine. New Engl. J. Medicine 383, 2603–2615, DOI: 10.1056/NEJMoa2034577 (2020). PMID: 33301246, https://doi.org/10.1056/NEJMoa2034577.

4. Voysey, M. et al. Safety and efficacy of the chadox1 ncov-19 vaccine (azd1222) against sars-cov-2: an interim analysis of four randomised controlled trials in brazil, south africa, and the uk. The Lancet 397, 99–111, DOI: 10.1016/S0140-6736(20)32661-1 (2021).

5. Baden, L. R. et al. Efficacy and safety of the mrna-1273 sars-cov-2 vaccine. New Engl. J. Medicine 384, 403–416, DOI: 10.1056/NEJMoa2035389 (2021). PMID: 33378609, https://doi.org/10.1056/NEJMoa2035389.

6. Pouwels, K. B. et al. Impact of delta on viral burden and vaccine effectiveness against new sars-cov-2 infections in the uk. medRxiv DOI: 10.1101/2021.08.18.21262237 (2021). https://www.medrxiv.org/content/early/2021/08/24/2021.08.18.21262237.full.pdf.

7. Lopez Bernal, J. et al. Effectiveness of covid-19 vaccines against the b.1.617.2 (delta) variant. New Engl. J. Medicine 385, 585–594, DOI: 10.1056/NEJMoa2108891 (2021). https://doi.org/10.1056/NEJMoa2108891.

8. Sheikh, A., McMenamin, J., Taylor, B. & Robertson, C. Sars-cov-2 delta voc in scotland: demographics, risk of hospital admission, and vaccine effectiveness. The Lancet 397, 2461–2462, DOI: 10.1016/S0140-6736(21)01358-1 (2021).

9. Imai, N. et al. Interpreting estimates of coronavirus disease 2019 (covid-19) vaccine efficacy and effectiveness to inform simulation studies of vaccine impact: a systematic review [version 1; peer review: awaiting peer review]. Wellcome Open Res. 6, DOI: 10.12688/wellcomeopenres.16992.1 (2021).

10. UK COVID-19 vaccines delivery plan. https://www.gov.uk/government/publications/uk-covid-19-vaccines-delivery-plan/uk-covid-19-vaccines-delivery-plan. Accessed: 2021-09-02.

11. All young people aged 16 and 17 in England to be offered vaccine by next week. https://www.gov.uk/government/news/all-young-people-aged-16-and-17-in-england-to-be-offered-vaccine-by-next-week. Accessed: 2021-09-02.

12. Universal vaccination of children and young people aged 12 to 15 years against covid-19. https://www.gov.uk/government/publications/universal-vaccination-of-children-and-young-people-aged-12-to-15-years-against-covid-19/universal-vaccination-of-children-and-young-people-aged-12-to-15-years-against-covid-19. Accessed: 2021-09-22.

13. COVID-19 vaccination programme for children and young people: guidance for schools. https://www.gov.uk/government/publications/covid-19-vaccination-resources-for-schools/covid-19-vaccination-programme-for-children-and-young-people-guidance-for-schools. Accessed: 2021-09-22.

14. Viner, R. M. et al. School closure and management practices during coronavirus outbreaks including covid-19: a rapid systematic review. The Lancet Child & Adolesc. Heal. 4, 397–404 (2020).

15. Viner, R. M. et al. Susceptibility to SARS-CoV-2 Infection Among Children and Adolescents Compared With Adults: A Systematic Review and Meta-analysis. JAMA Pediatr. 175, 143–156, DOI: 10.1001/jamapediatrics.2020.4573 (2021). https://jamanetwork.com/journals/jamapediatrics/articlepdf/2771181/jamapediatrics_viner_2020_oi_200071_1611604170.25358.pdf.

16. Keeling, M. J. et al. The impact of school reopening on the spread of covid-19 in england. Philos. Transactions Royal Soc. B 376, 20200261 (2021).

17. Panovska-Griffiths, J. et al. Determining the optimal strategy for reopening schools, the impact of test and trace interventions, and the risk of occurrence of a second covid-19 epidemic wave in the uk: a modelling study. The Lancet Child & Adolesc. Heal. 4 (2020).

18. Sonabend, R. et al. Non-pharmaceutical interventions, vaccination and the delta variant: epidemiological insights from modelling england’s covid-19 roadmap out of lockdown. medRxiv (2021).

19. Aylett-Bullock, J. et al. June: open-source individual-based epidemiology simulation. Royal Soc. Open Sci. 8, 210506, DOI: 10.1098/rsos.210506 (2021). https://royalsocietypublishing.org/doi/pdf/10.1098/rsos.210506.

20. Aylett-Bullock, J. et al. Operational response simulation tool for epidemics within refugee and idp settlements. medRxiv DOI: 10.1101/2021.01.27.21250611 (2021). https://www.medrxiv.org/content/early/2021/07/06/2021.01.27.21250611.full.pdf.

21. Chu, D. K. et al. Physical distancing, face masks, and eye protection to prevent person-to-person transmission of SARS-CoV-2 and COVID-19: a systematic review and meta-analysis. The Lancet (2020).

22. Wang, Y. et al. Reduction of secondary transmission of SARS-CoV-2 in households by face mask use, disinfection and social distancing: a cohort study in Beijing, China. BMJ Glob. Heal. 5, e002794 (2020).

23. Ahmed, F., Zviedrite, N. & Uzicanin, A. Effectiveness of workplace social distancing measures in reducing influenza transmission: a systematic review. BMC public health 18, 518 (2018).

24. Howard, J. et al. An evidence review of face masks against covid-19. Proc. Natl. Acad. Sci. 118, DOI: 10.1073/pnas.2014564118 (2021). https://www.pnas.org/content/118/4/e2014564118.full.pdf.

25. Fischer, E. P. et al. Low-cost measurement of face mask efficacy for filtering expelled droplets during speech. Sci. Adv. 6, eabd3083 (2020).

26. Feng, Y., Marchal, T., Sperry, T. & Yi, H. Influence of wind and relative humidity on the social distancing effectiveness to prevent COVID-19 airborne transmission: A numerical study. J. aerosol science 105585 (2020).

27. Atkinson, J. et al. Natural ventilation for infection control in health-care settings (World Health Organization, 2009).

28. Dai, H. & Zhao, B. Association of the infection probability of covid-19 with ventilation rates in confined spaces. Build Simul 1–7, DOI: 10.1007/s12273-020-0703-5 (2020).

29. Kerr, C. C. et al. Covasim: An agent-based model of covid-19 dynamics and interventions. PLoS Comput. Biol 7 (2021).

30. Di Domenico, L., Pullano, G., Sabbatini, C. E., Boëlle, P.-Y. & Colizza, V. Modelling safe protocols for reopening schools during the covid-19 pandemic in france. Nat. Commun. 12, 1073 (2021).

31. Head, J. R. et al. School closures reduced social mixing of children during covid-19 with implications for transmission risk and school reopening policies. J. The Royal Soc. Interface 18, 20200970, DOI: 10.1098/rsif.2020.0970 (2021). https://royalsocietypublishing.org/doi/pdf/10.1098/rsif.2020.0970.

32. Cohen, J. A., Mistry, D., Kerr, C. C. & Klein, D. J. Schools are not islands: Balancing covid-19 risk and educational benefits using structural and temporal countermeasures. medRxiv DOI: 10.1101/2020.09.08.20190942 (2020). https://www.medrxiv.org/content/early/2020/09/10/2020.09.08.20190942.full.pdf.

33. Courtemanche, C., Garuccio, J., Le, A., Pinkston, J. & Yelowitz, A. Strong social distancing measures in the united states reduced the covid-19 growth rate. Heal. Aff. 39, 1237–1246, DOI: 10.1377/hlthaff.2020.00608 (2020). PMID: 32407171, https://doi.org/10.1377/hlthaff.2020.00608.

34. Tatapudi, H. & Das, T. K. Impact of school reopening on pandemic spread: A case study using an agent-based model for covid-19. Infect. Dis. Model. 6, 839–847, DOI: https://doi.org/10.1016/j.idm.2021.06.007 (2021).

35. Rypdal, M. et al. Modelling suggests limited change in the reproduction number from reopening norwegian kindergartens and schools during the covid-19 pandemic. PLoS ONE 2 (2021).

36. Iwata, K., Doi, A. & Miyakoshi, C. Was school closure effective in mitigating coronavirus disease 2019 (covid-19)? time series analysis using bayesian inference. Int. J. Infect. Dis. 99, 57–61, DOI: https://doi.org/10.1016/j.ijid.2020.07.052 (2020).

37. Mishra, S. et al. Comparing the responses of the uk, sweden and denmark to covid-19 using counterfactual modelling. Sci. Reports 11, 16342, DOI: https://doi.org/10.1038/s41598-021-95699-9 (2021).

38. Knock, E. S. et al. Key epidemiological drivers and impact of interventions in the 2020 sars-cov-2 epidemic in england. Sci. Transl. Medicince 13 (2021).

39. People who have received vaccinations, by report date. https://coronavirus.data.gov.uk/details/vaccinations?areaType=nation&areaName=England. Accessed: 2021-09-02.

40. Public health england. investigation of novel sars-cov-2 variant: technical briefing1 (2021). https://assets.publishing.service.gov.uk/government/uploads/system/uploads/attachment_data/file/1005395/23_July_2021_Risk_assessment_for_SARS-CoV-2_variant_Delta.pdf. Accessed: 2021-08-23.

41. Gazit, S. et al. Comparing SARS-CoV-2 natural immunity to vaccine-induced immunity: reinfections versus breakthrough infections. medRxiv (2021).

42. Keeling, M. J. et al. Fitting to the uk covid-19 outbreak, short-term forecasts and estimating the reproductive number. medRxiv DOI: 10.1101/2020.08.04.20163782 (2021). https://www.medrxiv.org/content/early/2021/07/27/2020.08.04.20163782.full.pdf.

43. Dyson, L. et al. Possible future waves of sars-cov-2 infection generated by variants of concern with a range of characteristics. medRxiv DOI: 10.1101/2021.06.07.21258476 (2021). https://www.medrxiv.org/content/early/2021/08/25/2021.06.07.21258476.full.pdf.

44. Coronavirus (covid-19) infection survey, uk: 16 july 2021. https://www.ons.gov.uk/peoplepopulationandcommunity/healthandsocialcare/conditionsanddiseases/bulletins/coronaviruscovid19infectionsurveypilot/16july2021. Accessed: 2021-08-23.

45. Coronavirus details on vaccinations for england. https://coronavirus.data.gov.uk/details/vaccinations?areaType=nation&areaName=England. Accessed: 2021-08-10.

46. COVID-19 in Scotland. https://public.tableau.com/app/profile/phs.covid.19/viz/COVID-19DailyDashboard_15960160643010/Overview. Accessed: 2021-09-02.

47. Craig, P. S., Goldstein, M., Seheult, A. H. & Smith, J. A. Pressure matching for hydrocarbon reservoirs: a case study in the use of bayes linear strategies for large computer experiments (with discussion). In Gatsonis, C. et al. (eds.) Case Studies in Bayesian Statistics, vol. 3, 36–93 (SV, New York, 1997).

48. Vernon, I., Goldstein, M. & Bower, R. G. Galaxy formation: a bayesian uncertainty analysis. Bayesian Analysis 5, 619–670 (2010).

49. Andrianakis, I. et al. Bayesian history matching of complex infectious disease models using emulation: A tutorial and a case study on HIV in uganda. PLoS Comput. Biol. 11, e1003968 (2015).

50. Read, J. M. et al. Hospital-acquired SARS-CoV-2 infection in the UK’s first COVID-19 pandemic wave. The Lancet 398, 1037–1038, DOI: 10.1016/S0140-6736(21)01786-4 (2021).

51. Coronavirus (COVID-19) latest insights: Lifestyle. https://www.ons.gov.uk/peoplepopulationandcommunity/healthandsocialcare/conditionsanddiseases/articles/coronaviruscovid19latestinsights/lifestyle#compliance-with-covid-19-guidance. Accessed: 2021-09-05.

52. Self-isolation removed for double-jabbed close contacts from 16 August. https://www.gov.uk/government/news/self-isolation-removed-for-double-jabbed-close-contacts-from-16-august. Accessed: 2021-09-02.

53. YouGov COVID-19 behaviour changes tracker: Avoiding going to work. https://yougov.co.uk/topics/international/articles-reports/2020/03/17/personal-measures-taken-avoid-covid-19 (2021).

54. Coronavirus Job Retention Scheme statistics: 29 July 2021. https://www.gov.uk/government/statistics/coronavirus-job-retention-scheme-statistics-29-july-2021/coronavirus-job-retention-scheme-statistics-29-july-2021. Accessed: 2021-09-05.

55. Coronavirus and key workers in the uk. https://www.ons.gov.uk/employmentandlabourmarket/peopleinwork/earningsandworkinghours/articles/coronavirusandkeyworkersintheuk/2020-05-15 (2020).

56. YouGov COVID-19 behaviour changes tracker: Wearing a face mask when in public places. https://yougov.co.uk/topics/international/articles-reports/2020/03/17/personal-measures-taken-avoid-covid-19 (2021).

57. The state of the restaurant industry. https://www.opentable.com/state-of-industry (2021).

58. Quera-Bofarull, A. et al. JUNE 1.1.1: School reopening paper. Zenodo/Github, DOI: 10.5281/zenodo.5521173 (2021).

59. Hill, N. et al. 2020 government greenhouse gas conversion factors for company reporting: Methodology paper for conversion factors final report (2020).

60. Deaths within 28 days of positive test by date of death. https://coronavirus.data.gov.uk/details/deaths?areaType=nation&areaName=England. Accessed: 2021-09-02.

61. SPI-M-O: Consensus statement on COVID-19, 30 June 2021.

62. Volz, E. et al. Transmission of SARS-CoV-2 Lineage B.1.1.7 in England: Insights from linking epidemiological and genetic data. Tech. Rep. (2021). DOI: 10.1101/2020.12.30.20249034. Company: Cold Spring Harbor Laboratory Press Distributor: Cold Spring Harbor Laboratory Press Label: Cold Spring Harbor Laboratory Press Type: article.

